# A preliminary report on alcohol-associated DNA methylation and suicidal behavior: evidence using Mendelian randomization

**DOI:** 10.1101/19013300

**Authors:** Charleen D. Adams

## Abstract

Alcohol consumption, a risk factor for suicide, is associated with changes in DNA methylation, which may play a role in the pathophysiology of suicidality. Here, Mendelian randomization probes whether alcohol-associated changes in DNA methylation mediate risk for suicidal behavior. The results suggest that higher alcohol-associated DNA methylation levels at cg18120259 confer a weak causal effect. cg18120259 is instrumented by a cis-methylation quantitative trait locus (rs9472155) that is upstream of *VEGFA* and associated with circulating VEFG levels. To ascertain whether the cg18120259 finding is confounded by VEGF levels, a Mendelian randomization of VEGF levels on suicidality was performed, revealing a null effect. This implies circulating VEGF levels (observed among suicide attempters) may mark but not cause suicide. Both mechanisms should be re-examined in large replication studies, as the information might be harnessed for early detection of at-risk individuals and possibly save lives.

---

Alcohol consumption is a strong risk factor for suicide^1^ and is associated with changes in DNA methylation^2^ and chronic inflammation^3^, both of which may play a role in the pathophysiology of suicidality. Lower plasma concentrations of vascular endothelial growth factor (VEGF) have been documented for suicide attempters versus controls^4^. The present paper uses Mendelian randomization (MR) to investigate whether alcohol-associated changes in DNA methylation cause suicidal behavior and considers possibilities that could explain the relationships between changes in DNA methylation, circulating VEGF levels, and suicidality (used throughout to denote attempts and completions, not suicidal ideation): vertical pleiotropy, horizontal pleiotropy, and confounding pleiotropy (Fig. 1.).

**Fig. 1.**
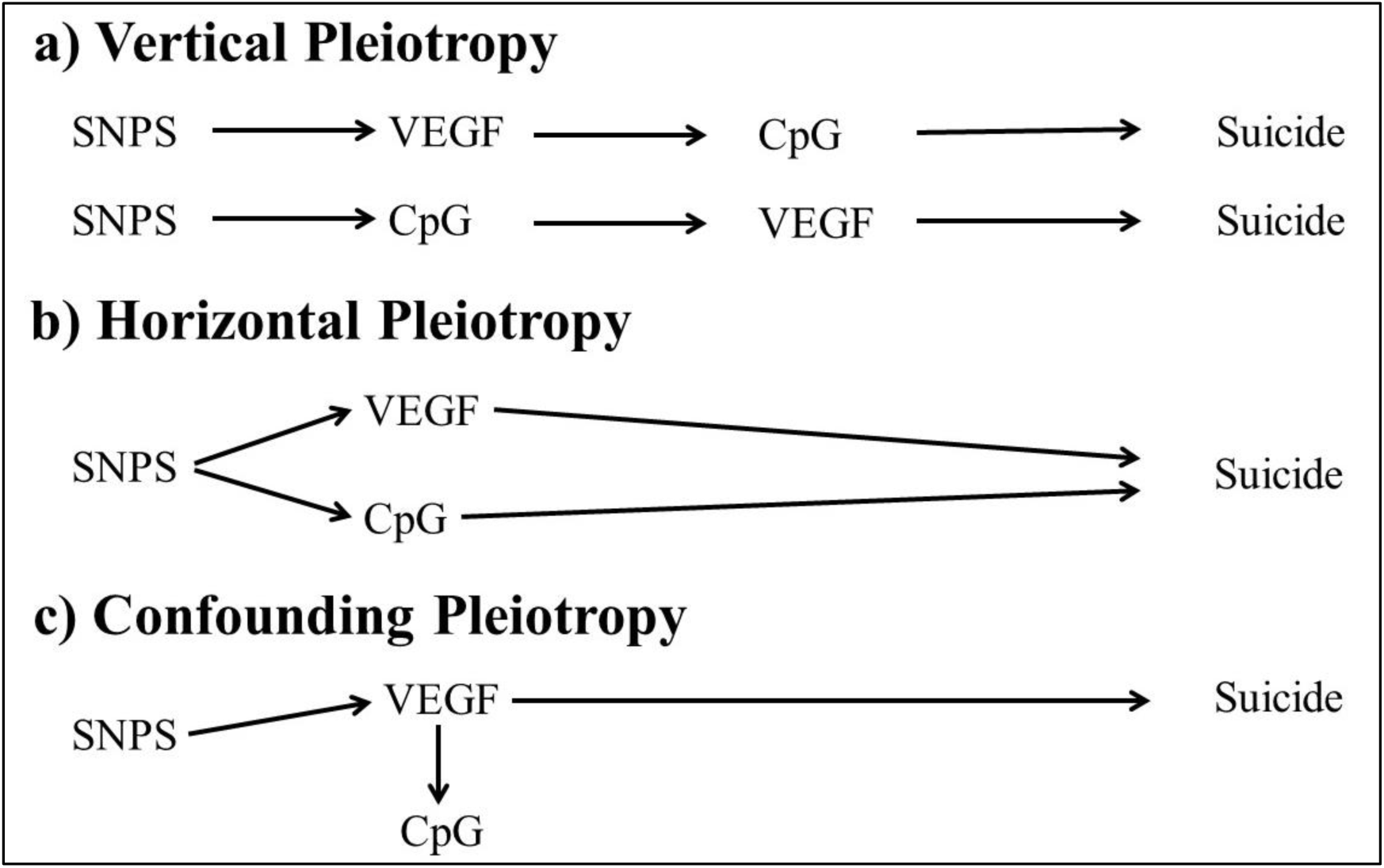
Possible pathways that might explain pleiotropy between DNA methylation, VEGF levels, and suicidal behavior. The pathways depicted here do not preclude confounding by unmeasured traits. (“CpG” denotes a site in the genome where a cytosine nucleotide is followed by a guanine nucleotide in the linear sequence of bases. CpGs can be methylated and their levels measured.)

## Conceptual framework

MR is an instrumental variables technique, analogous to a randomized-controlled trial. It uses genetic variants (typically single-nucleotide polymorphisms, SNPs) strongly associated with traits in statistical models instead of traits themselves. Doing so avoids most environmental sources of confounding and averts reverse causation^5–7^. Two-sample MR is an extension of the procedure that uses summary statistics from two genome-wide association (GWA) studies^8–13^. For example, the causal effect of DNA (CpG) methylation on suicidality can be assessed as follows (Fig. 2): Estimates of the SNP-CpG associations 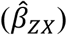 are calculated in Sample 1, a GWA study of DNA methylation levels (SNPs strongly associated with CpG levels are termed methylation quantitative trait loci (mQTL)). The association between these same SNPs and suicidality 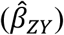 are then estimated in a GWA study of a suicidality phenotype (Sample 2). These estimates are combined into Wald ratios 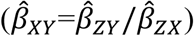 to produce a causal estimate of CpG methylation on suicidality.

**Fig. 2.**
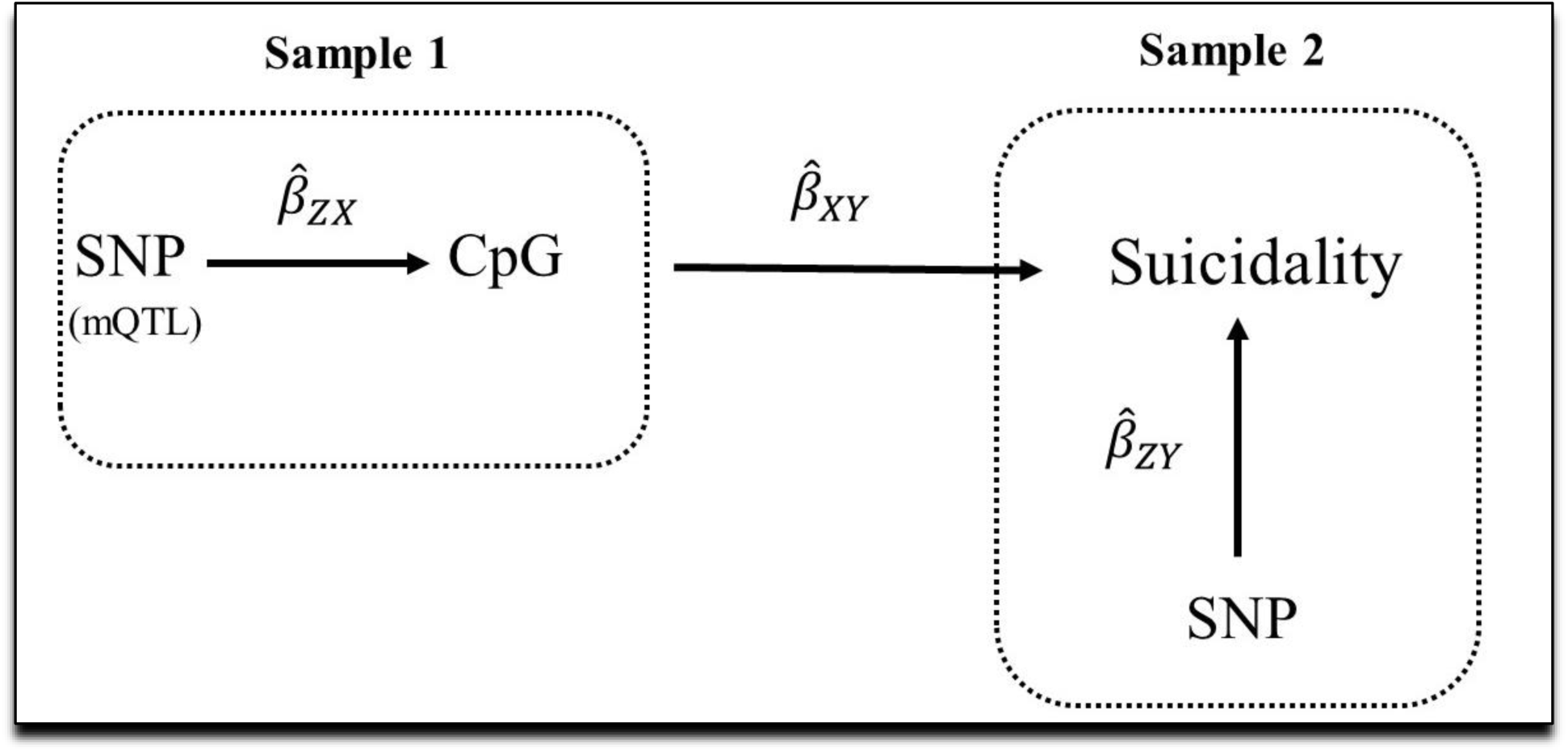
Two-sample MR depicting the use of methylation quantitative trait loci (mQTL) to instrument DNA methylation (CpG) levels (Sample 1) to test the effect of alcohol-associated DNA methylation changes on suicidality. mQTL identified in Sample 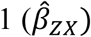 (are extracted from Sample 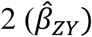. The effect estimates are combined into Wald ratios 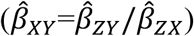.

## Mendelian randomization assumptions

Three assumptions must hold for MR to be valid: (i) the SNPs acting as the instrumental variables must be strongly associated with the exposure; (ii) the instrumental variables must be independent of confounders of the exposure and the outcome; and (iii) the instrumental variables must be associated with the outcome only through the exposure^10,14^.

## Results

Higher alcohol-associated methylation at cg18120259 is suggestively weakly causal for suicide (OR per beta-level increase in DNA methylation: 1.0005; 95% CI 1.0002, 1.0008; *P*=0.001). These findings replicated with a sensitivity test using a closely related ICD-9 outcome GWA study for suicidality (OR per beta-level increase in DNA methylation: IVW: 1.0003; 95% CI 1.00003, 1.0007; *P*=0.03). None of the seven remaining CpGs showed evidence for causality (Table 1).

**Table 1.** Causal effect estimates for the association of alcohol-associated DNA methylation on risk for suicide.

| CpG | Suicide phenotype | MR method | Num. SNPs | Odds Ratio | Lower 95% CI | Upper 95% CI | P-value |
| --- | --- | --- | --- | --- | --- | --- | --- |
| cg18120259 | ICD-9 E9500 | Wald ratio | 1 | 1.0005 | 1.0002 | 1.0008 | 0.001 |
| cg18120259 | ICD-9 E9503* | Wald ratio | 1 | 1.0003 | 1.0000 | 1.0006 | 0.030 |
| cg02583484 | ICD-9 E9500 | Wald ratio | 1 | 1.0000 | 0.9996 | 1.0003 | 0.866 |
| cg02583484 | ICD-9 E9503* | Wald ratio | 1 | 0.9998 | 0.9994 | 1.0001 | 0.155 |
| cg07626482 | ICD-9 E9500 | Wald ratio | 1 | 0.9999 | 0.9996 | 1.0001 | 0.312 |
| cg07626482 | ICD-9 E9503 | Wald ratio | 1 | 1.0000 | 0.9997 | 1.0003 | 0.973 |
| cg14476101 | ICD-9 E9500 | IVW | 2 | 1.0000 | 0.9999 | 1.0001 | 0.663 |
| cg14476101 | ICD-9 E9503* | IVW | 2 | 1.0000 | 0.9999 | 1.0001 | 0.519 |
| cg16246545 | ICD-9 E9500 | Wald ratio | 1 | 1.0000 | 0.9998 | 1.0001 | 0.484 |
| cg16246545 | ICD-9 E9503* | Wald ratio | 1 | 1.0000 | 0.9999 | 1.0001 | 0.770 |
| cg26170244 | ICD-9 E9500 | Wald ratio | 1 | 1.0000 | 0.9996 | 1.0004 | 0.916 |
| cg26170244 | ICD-9 E9503* | Wald ratio | 1 | 1.0001 | 0.9997 | 1.0005 | 0.670 |
| cg11701312 | ICD-9 E9500 | Wald ratio | 1 | 1.0002 | 0.9999 | 1.0005 | 0.235 |
| cg11701312 | ICD-9 E9503* | Wald ratio | 1 | 0.9999 | 0.9996 | 1.0002 | 0.654 |
| cg06469895 | ICD-9 E9500 | Wald ratio | 1 | 1.0001 | 0.9997 | 1.0004 | 0.674 |
| cg06469895 | ICD-9 E9503* | Wald ratio | 1 | 0.9998 | 0.9995 | 1.0001 | 0.255 |

\*Denotes a sensitivity replication. IVW=inverse-weighted variance. CI=confidence interval.

### Sensitivity analyses

The sensitivity analysis of circulating VEGF levels on risk for suicide was null (Table 2): IVW estimate 1.0000, 95% CI 0.9999, 1.0001; *P*=0.6776). The sensitivity analysis for frequency of alcohol consumed with meals on risk for suicide demonstrates a causal effect for more drinks consumed (OR for suicide per category higher of drinking: IVW estimate 1.0013; 95% CI 1.0002, 1.0024; *P*=0.0184).

**Table 2.** Causal effect estimates for VEGF levels on risk for suicide.

| MR test | SNPs | Odds ratio | Lower 95% CI | Upper 95% CI | P-value |
| --- | --- | --- | --- | --- | --- |
| IVW | 3 | 1.0000 | 0.9999 | 1.0001 | 0.6776 |
| MR-Egger | 3 | 1.0000 | 0.9998 | 1.0001 | 0.7416 |
| Weighted median | 3 | 1.0000 | 0.9999 | 1.0001 | 0.8373 |
| Weighted mode | 3 | 1.0000 | 0.9999 | 1.0001 | 0.7791 |

IVW=inverse-variance weighted. CI=confidence interval.

## Discussion

Regarding the relationships between DNA methylation, VEGF levels, and suicidality, four possibilities are considered: (i) VEGF levels could influence suicide through cg18120259 (vertical pleiotropy); (ii) cg18120259 could regulate VEGF levels and hence suicidality (vertical pleiotropy); (iii) cg18120259 and VEGF levels could have independent, direct influences on suicidality (horizontal pleiotropy); (iv) and the findings implicating methylation of cg18120259 in suicidality could be explained by confounding from an unmeasured trait; i.e., the hypermethylation of cg18120259 could be a biomarker associated with a causal trait in individuals who are at-risk for suicide.

cg18120259 is instrumented by a cis-mQTL (rs9472155) that is associated with VEGF levels. Since higher methylation levels typically downregulate expression, these findings might imply that alcohol-associated higher methylation of cg18120259 acts to downregulate *VEGFA*. However, from the MR results, there is no evidence for a direct, causal effect of VEGF levels on risk for suicide, suggesting the lower levels of VEGF that have been previously reported in suicide attempters are marking but not causing the behavior. This is evidence against explanations (i), (ii), and (iii); i.e., the finding for hypermethylation of cg18120259 increasing risk for suicide does not appear to be confounded by VEGF levels or occurring through them.

Because VEGF levels are the only documented trait associated with rs9472155 (Supplementary Table 15), the null MR provides some evidence against horizontal pleiotropy. Notwithstanding, the numbers of suicide cases in the two outcome GWA are small. Exploring the possibility that VEGF confers a causal effect should be reconsidered when more cases are available in public GWA studies of suicide. Further, the small number of suicide cases for the tests of DNA methylation on suicidality might also have led to Type 1 error. This possibility is mitigated some by the replication with the MR analysis of the closely related suicidality phenotype and by the MR of frequency of alcohol consumed with meals on suicidality, which showed that frequency of drinking among drinkers does increase risk for suicide. It is unlikely that these separate and biologically plausible tests performed with three separate outcome GWA studies of suicidality are chance artefacts.

That leaves confounding by an unmeasured endophenotype (iv). A look-up of rs9472155 in the UCSC Genome Browser reveals that it is at transcription factor binding site for the estrogen receptor alpha (ERα), encoded by *ESR1* (6q25.1)^15^. However, while estrogen has been hypothesized to play a role in the pathophysiology of suicidality, a case-control study examining genetic variants in *ESR1* among suicide attempters and healthy controls found no evidence that ERα contributes strongly to the risk for suicide^16^. Thus, while a limitation of the present study is that computational methods to test pleiotropy directly were unavailable for investigation of DNA methylation, probing biologically plausible confounders revealed little evidence for confounding (iv). Since the possibility cannot be ruled out, hypermethylation of cg18120259 could be a biomarker that tags a true underlying causal factor or itself a molecular driver of alcohol-associated suicide. Either way, should these preliminary findings replicate in future studies, this information might be harnessed for early detection of at-risk individuals, thereby possibly reducing the burden of suicide – it could help save lives.

A strength of this study is that there is unlikely to be much participant overlap between the exposure and outcome GWA studies used in the primary two-sample MR designs^17^. This means that bias from sample overlap is minimized.

Future MR investigations of alcohol-associated DNA methylation changes on suicidality would best be done using multivariable MR – with full summary data for alcohol-induced DNA methylation and full summary data of VEGF levels (currently not available publicly) – and include an outcome GWA of suicidality that includes more cases. Multivariable MR would permit a formal statistical assessment of the pleiotropy between DNA methylation and VEGF. Though the sensitivity analyses here already do this, including more cases would reduce concerns for both Type 1 and 2 error from a possible lack of statistical power.

## Methods

70 CpGs associated with alcohol consumption in 1,135 European American male veterans were extracted from the manuscript by Xu et al. (2019) (Supplementary Table 3)^2^. To determine whether the CpGs were associated with cis-methylation quantitative trait loci (mQTLs), the list of 70 CpGs were uploaded to the mQTL database (http://www.mqtldb.org/)^18^, revealing that 8/70 CpGs have cis-mQTLs associated with them at genome-wide significance (*P* < 5 x 10-8) (Supplementary Tables 4-5). Trans-mQTLs were excluded. Summary statistics for these eight CpGs functioned as data sources for SNP-CpG associations and used in two-sample Mendelian randomization (MR). For each of the eight CpG instruments, only cis-mQTLs that were independent (those not in linkage disequilibrium, LD; R^2^ < 0.01) were kept.

A genome-wide association (GWA) study performed by the Neale lab^19^ on UK Biobank data field 41205^20,21^ for ICD-9 code E9500 (“suicide, self-inflicted poisoning by analgesics, antipyretics, antirheumatics) was accessed via MR-Base (http://www.mrbase.org/)^8^ and served as the outcome data source for the SNP-suicidality associations (sample size=462,859 of which 151 were cases). Harmonized SNP-CpG and SNP-outcome associations were analyzed as Wald ratios or combined with the inverse-variance weighted (IVW) method when more than one SNP instrumented a CpG site (Fig. 2). This was done within the “TwoSampleMR” package^8^. To correct for running eight tests, a Bonferroni threshold was set to P<0.006 (0.05/8). Analyses were performed in in R version 3.5.2. All data sources used for SNP-CpG and SNP-suicidality associations are publicly available.

### Sensitivity analyses

None of the DNA methylation instruments contained a sufficient number of SNPs to perform typical MR sensitivity analyses for pleiotropy, such as MR-Egger regression. To address this, a second two-sample MR was run as a replication for each instrument using a different UK Biobank ICD-9 code for a closely related suicidality phenotype (E9503: “suicide and self-inflicted poisoning by tranquillizers, other psychotropic agents”; sample size=462,569 of which 141 are cases). In addition, PhenoScanner, a curated database of GWA studies containing SNP-phenotype associations, was used to ascertain potential pleiotropic confounders for the SNP instrument for cg18120259 (rs9472155)^22,23^. PhenoScanner revealed that the SNP instrumenting cg18120259 (rs9472155) is associated with VEGF, altered levels of which have been documented in those who attempt suicide. Therefore, to investigate the possibility that alcohol-induced DNA methylation at cg18120259 is confounded by the influence of VEGF, an MR of VEGF levels on suicidality was performed as a sensitivity test. For this, an instrument for circulating VEGF levels was constructed from a GWA study of 3,527 individuals of European descent in the Framingham Heart Study^24^.

The number of cases of suicide are small. To probe the possibility that outcome GWA with few cases of suicide could be used with biologically probable hypotheses, a final sensitivity test was performed among those who consume alcohol. In the UK Biobank, those who indicated they consumed alcohol were asked how frequently they drank with meals (data field 1618; sample size 172,454). An MR of frequency of drinking with meals on suicidality was run using a GWA for ICD-9 code E9509: “Suicide and self-inflicted poisoning by other/unspecified solid and liquid substances” (the primary UK Biobank ICD-9 code E9509 GWA was performed by the Neale lab^19^).

The same methods for instrument construction listed above for the DNA methylation instruments were used to instrument VEGF levels and frequency of alcohol consumed with meals.

### Instrument strength

*F*-statistics, which provide a measure of instrument strength, and R^2^ statistics (how much variance in a trait is explained by an instrument)^25^ were calculated for each primary test (Table 3). *F*-statistics <10 are weak^26^. All instruments used in this analysis are aptly strong. Detailed characteristics of proxies for all instruments are available in Supplementary Tables 6-14, and 16.

**Table 3.** Causal effect estimates for frequency of alcohol consumed with meals among those who drink on risk for suicide.

| MR test | SNPs | Odds ratio | Lower 95% CI | Upper 95% CI | P-value |
| --- | --- | --- | --- | --- | --- |
| IVW | 14 | 1.0013 | 1.0002 | 1.0024 | 0.0184 |
| MR-Egger | 14 | 1.0052 | 0.9943 | 1.0163 | 0.3694 |
| Weighted median | 14 | 1.0008 | 0.9994 | 1.0021 | 0.2783 |
| Weighted mode | 14 | 1.0004 | 0.9981 | 1.0026 | 0.7633 |

IVW=inverse-variance weighted. CI=confidence interval.

**Table 4.** Instrument strength.

| Trait | $R^2$ | *F*-statistic |
| --- | --- | --- |
| cg18120259 | 0.03 | 32 |
| cg02583484 | 0.03 | 35 |
| cg07626482 | 0.04 | 42 |
| cg14476101 | 0.18 | 252 |
| cg16246545 | 0.15 | 199 |
| cg26170244 | 0.01 | 17 |
| cg11701312 | 0.03 | 35 |
| cg06469895 | 0.02 | 27 |
| VEGF | 0.40 | 794 |
| Frequency of drinks with meals | 0.003 | 38 |

### Statistical software

All described analyses were performed in R version 3.5.2.

### Data availability

All data sources are publicly available. The data for the Xu *et al*. (2019) instrument of alcohol-associated DNA methylation levels^2^ and the data for the VEGF instrument^24^ were obtained directly from the supplementary files accompanying their primary papers. The remaining data used for these analyses are accessible within MR-Base http://www.mrbase.org/^8^. Template code for performing these analyses is also available through the MR-Base app.

## Data Availability

All data sources are publicly available. The data for the Xu et al. (2019) instrument of alcohol-associated DNA methylation levels and the data for the VEGF instrument were obtained directly from the supplementary files accompanying their primary papers. The remaining data used for these analyses are accessible within MR-Base.

http://www.mrbase.org/

## Acknowledgements

Thank you to the cohorts that made their GWA study summary data public.

## Additional information

Supplementary information accompanies this paper.

